# Disease Spectrum of Breast Cancer Susceptibility Genes

**DOI:** 10.1101/2020.08.11.20172007

**Authors:** Jin Wang, Preeti Singh, Kanhua Yin, Jingan Zhou, Yujia Bao, Menghua Wu, Kush Pathak, Sophia K. McKinley, Danielle Braun, Kevin S. Hughes

## Abstract

**Background:** Pathogenic variants in cancer susceptibility genes can increase the risk of a spectrum of diseases. We aim to evaluate the disease spectrum of breast cancer susceptibility genes (BCSGs) to develop a comprehensive resource of gene-disease associations for clinicians.

**Methods:** Thirteen genes (ATM, BARD1, BRCA1, BRCA2, CDH1, CHEK2, NBN, NF1, PALB2, PTEN, RECQL, STK11 and TP53), that have been conclusively established as BCSGs by the Clinical Genome Resource (ClinGen) and the NCCN guidelines, were investigated. For these thirteen genes, potential gene-disease associations were identified and evaluated based on six genetic resources (ClinGen, NCCN, OMIM, Genetics Home Reference, GeneCards and Gene-NCBI) and an additional literature review using a semiautomated natural language processing (NLP) abstract classification procedure.

**Results:** A total of 40 diseases were confirmed as being associated with one or more of the 13 BCSGs by our evaluation. Malignant diseases including prostate cancer, pancreatic cancer, colorectal cancer, brain tumor, gastric cancer, ovarian cancer, and sarcoma were associated with at least 3 BCSGs. Furthermore, a total of 87 gene-disease associations were confirmed by our evaluation, of which 85% (74/87) were confirmed by ClinGen and/or NCCN. Conversely, 9 gene-disease associations absent from both ClinGen and NCCN were confirmed in the other four genetic resources (≥3) and 4 gene-disease associations were confirmed by the NLP-based procedure.

**Conclusion:** This is the first study to systematically investigate the reported disease spectrum of BCSGs in multiple sources. Our innovative approach provides a general guide for evaluating gene-disease associations, and improves the clinical management for at-risk individuals.

## Introduction

Hereditary predisposition is found in approximately 10% of all breast cancer cases [1]. Most are related to germline mutations in high-penetrance genes like BRCA1 and BRCA2 [2-5]. Since the identification of BRCA1 and BRCA2 [6,7], genetic testing has become a routine part of clinical care for individuals with possible hereditary breast cancer predisposition [1]. With the substantial increase in knowledge of cancer genetics [8,9], more than 30 breast cancer susceptibility genes (BCSGs) have been suggested, including genes with high (e.g., *BRCA1/2, TP53, CDH1, PTEN*, and *STK11*), moderate (e.g., *PALB2, CHEK2, ATM*, and *RECQL*), and low-to-disputed penetrance (e.g., *MLH1, MSH2, MSH6, PMS2, MEN1*, and *PPM1D*) [9-12]. Among them, 13 genes with high or moderate penetrance for breast cancer have been definitively established by the Clinical Genome Resource (ClinGen) [11] and the National Comprehensive Cancer Network (NCCN) [12], the top two authoritative resources.

Pathogenic variants in a BCSG can also increase the risk of other diseases. For instance, *CDH1* is not only associated with increased breast cancer risk, but also a predisposition to gastric cancer [13,14]. Furthermore, several BCSGs are responsible for rare hereditary cancer syndromes, such as *TP53*, which is responsible for Li-Fraumeni syndrome. Individuals with this syndrome have a very high risk of developing multiple malignancies, including but not limited to, breast cancer, sarcoma, brain cancer, leukemia, lung cancer, and adrenocortical cancer [15-18]. As comprehensive panel genetic testing becomes the norm [19], clinicians are increasingly faced with the challenge of advising mutation carriers about genes they may be less familiar with or involving cancer susceptibility in organs outside their specialty.

A variety of resources, in addition to NCCN and ClinGen, describe the diseases associated with each gene [20], including but not limited to, Genetics Home Reference (https://ghr.nlm.nih.gov/), Online Mendelian Inheritance in Man (OMIM) (https://www.ncbi.nlm.nih.gov/omim), GeneCards (https://www.genecards.org/) and Gene-NCBI (https://www.ncbi.nlm.nih.gov/gene/). However, the gene-disease associations described among these six resources are often ambiguous, incomplete, or confusing. For example, the association of *BRCA2* with melanoma is identified in NCCN and Genetics Home Reference but not in other genetic resources such as ClinGen, OMIM, GeneCards, or Gene Reviews. Furthermore, some gene-disease associations are not found in any genetic resource, such as the association of *CHEK2* with gastric cancer, which has been established with high likelihood in the literature [21,22]. This poses a considerable dilemma for clinicians who are obligated to identify and assess gene-disease associations that require management in clinical practice. Relying on a patchwork of resources is cumbersome, time-consuming, and can lead to errors of omission. A single comprehensive resource is critically needed to streamline this process. In light of these issues, we have developed a novel approach to identify, evaluate, and curate the diseases or complex syndromes associated with cancer susceptibility genes, and we apply it to BCSGs in this study.

## Methods

1. **Established breast cancer susceptibility genes** Only monoallelic BCSGs were included in the present study. The BCSGs were initially identified using ClinGen [11] and NCCN [12]. In 2019, Lee et al., along with other experts on the ClinGen Hereditary Cancer Clinical Domain Executive Committee, published a list of 31 high-priority genes for curation using the ClinGen Gene Curation clinical validity framework [11]. Among them, 11 genes were classified as having a ‘Definitive’ or ‘Moderate’ association with breast cancer and these were included in our study. The NCCN Guidelines for ‘Genetic/Familial High-Risk Assessment: Breast and Ovarian’ identified 20 genes offered in multi-gene panels where breast cancer risk was classified as ‘Increase’, ‘Potential increase’, or ‘Unknown/Insufficient evidence’ [12]. Of these 20, the 12 genes which were classified as ‘Increase’ or ‘Potential increase’ were also included in our study. Accounting for overlap between these two resources, 13 BCSGs were selected for breast cancer, namely, ATM, BARD1, BRCA1, BRCA2, CDH1, CHEK2, NBN, NF1, PALB2, PTEN, RECQL, STK11, and TP53 (**Figure 1**).
2. **Identification of gene-disease association** Diseases associated with BCSGs were initially identified in the six genetic resources (ClinGen, NCCN, OMIM, Genetics Home Reference, GeneCards, and Gene-NCBI) and by reviewing the literature. For each of these sources, each potential association was coded in our database as ‘1’ if the association was definitive, ‘9’ if the association was possible, and ‘0’ if there was no association, as shown in **Supplementary Table 1**. The date of last access to all resources was January 8, 2020. In the following sections we describe in detail each of these resources.

### 2.1. ClinGen

ClinGen is a database curated by the Clinical Genome Resource, which uses a standardized clinical validity framework to assess evidence to validate a gene-disease association and to define disease management. We extracted data regarding gene-disease associations directly from the ‘Gene-Disease Validity’ reports in ClinGen (https://search.clinicalgenome.org/kb/gene-validity).

The strength of ‘Gene-Disease Validity’ was classified by ClinGen as ‘Definitive’, ‘Strong’, ‘Moderate’, ‘Limited’, ‘Refuted’, ‘Disputed’, or ‘No Reported Evidence’ based on the level of evidence. If an association was classified as ‘Definitive’, ‘Strong’, or ‘Moderate’, it was coded in our database as ‘1’ in the field ClinGen Validity. If an association was classified as ‘Limited’, it was coded in our database as ‘9’. If an association was classified as ‘Refuted’, ‘Disputed’ or ‘No Reported Evidence’, it was coded in our database as ‘0’.

We also reviewed the ‘Actionability’ reports in ClinGen, where the gene-disease associations were identified indirectly (https://clinicalgenome.org/working-groups/actionability/). The ‘Actionability’ report in ClinGen summarizes secondary findings in patients and identifies diseases caused by susceptibility genes that can be prevented or palliated. A gene-disease association was coded as ‘1’ in our database in the field ClinGen Actionability, if the disease was a manifestation of the genetic disorder, if management of that disease was recommended by screening or preventive intervention, or if the disease was confirmed in the ‘Penetrance’ section of the ‘Actionability’ report. The gene-disease association was coded in our database as ‘9’, if the report suggested a possible relationship.

### 2.2. NCCN Guidelines

Data was extracted from the NCCN Guidelines® Genetic/Familial High-Risk Assessment: Breast and Ovarian Version 3.2019 [12] and Colorectal Version 2.2019 [23]. A gene-disease association was coded as ‘1’ in our database, if a disease or a feature was used to identify patients for genetic testing or if the management of a disease was recommended for mutation carriers. If NCCN identified a possible relationship, the gene-disease association was coded in our database as ‘9’.

### 2.3. Other Genetic Resources

Other reputable databases such as ‘OMIM’, ‘Genetics Home Reference’, ‘GeneCards’, and ‘Gene-NCBI’ (described in detail below) were also used to identify the gene-disease associations. If a gene-disease association was present in a resource, this association was coded as ‘1’ in our database.

‘OMIM’ is an online compendium of human genes and genetic phenotypes that is written and regularly updated by the McKusick-Nathans Institute of Genetic Medicine. The “Clinical Synopses” table for each gene was used to identify gene-disease associations.

‘Genetics Home Reference’ is a free online resource that was created after the announcement of the human genome map in 2003 and is maintained by the National Library of Medicine. It is designed to make the connection between genetics and disease more transparent for the general public. The “health conditions related to the Genetic Changes” section for each gene was used to identify gene-disease associations.

‘GeneCards’ is a comprehensive database of human genes. The content of this database is reviewed and updated by the GeneCards Suite Project Team. The “disorders” table for each gene was used to identify gene-disease associations.

‘Gene-NCBI’ is a resource of the National Center for Biotechnology Information (NCBI), which centralizes gene-related information into individual records. Many different types of gene-specific data are connected to the record including gene products and their attributes, expression, interactions, pathways, variation and its phenotypic consequences. The “Phenotypes” section for each gene was used to identify gene-disease associations.

## 3. Evaluation of gene-disease association

The process of validating the gene-disease association is outlined in **Figure 1**. Of the six genetic resources, we considered ClinGen and NCCN the most authoritative and curated these as major resources. As shown in **Figure 1**, we assigned the gene-disease association ‘confirmed’ if it was coded as ‘1’ in either ClinGen or NCCN. Additionally, if the gene-disease association was coded as ‘1’ in ≥3 other genetic resources (OMIM, Genetic Home Reference, GeneCard, and Gene-NCBI), it was also assigned ‘confirmed’. On the other hand, we assigned the gene-disease association ‘uncertain’, if it was not coded as ‘1’ in either ClinGen or NCCN, and in <3 of the other genetic resources (OMIM, Genetic Home Reference, GeneCard, and Gene-NCBI). We assigned the gene-disease association ‘no association’ directly, if it was coded as ‘0’ in ClinGen.

All ‘uncertain’ gene-disease associations were further evaluated by literature review using an abstract classifier NLP procedure, which classifies abstract as being relevant to cancer penetrance or not [24,25]. Our NLP abstract classifier was developed to cull germline penetrance papers from PubMed. In brief, it uses a Support Vector Machine algorithm to classify abstracts as relevant to penetrance, prevalence, both, or neither [25]. This NLP abstract classifier has been incorporated into a semiautomated procedure. The sensitivity and specificity of this approach in identifying cancer penetrance studies have been validated [24].

In this study, we used standard gene and disease PubMed search terms (**Supplementary Table 2**) to run the procedure. The NLP abstract classifier was applied to identify the abstracts that were classified as relevant to prevalence or penetrance, which were subsequently reviewed by two researchers independently. We then retrieved the full text of these penetrance studies and determined the gene-disease associations based on the quality of the penetrance study (including type of study, sample size, carrier numbers, and ascertainment criteria) as well as the statistical significance of the results.

If no relevant penetrance abstract was identified, the association was assigned as ‘no association’. If relevant penetrance studies were identified, all of them were presented in a group meeting in which the PI (KSH) and 4-5 research fellows (MDs) participated. The attendees selected high-quality penetrance studies based on study design, patient population, number of pathogenic variant carriers, and ascertainment mechanism, and reached a final consensus based on evaluating these high-quality studies. One rule of thumb was we considered a gene-cancer association to be real if at least one high-quality penetrance study reported at least 2-fold increased risk that was statistically significant. If the attendees could not reach a consensus, the gene-disease association remained ‘uncertain’. Of note, to ensure accuracy, the group meeting not only discussed the potential controversial gene-cancer associations but also examined all the evidence regarding every gene-cancer associations reported in the study.

## Results

### 1. Breast cancer susceptibility genes in 6 genetic resources

As shown in **Table 1**, among the thirteen established BCSGs, the association of breast cancer risk with *ATM, BARD1, BRCA1, BRCA2, CDH1*, and *CHEK2* was identified in all six genetic sources; *NBN, PALB2, PTEN, STK11* and *TP53* were identified in at least two genetic sources. However, the association of breast cancer risk with *NF1* was only identified in NCCN, and *RECQL* was only identified in ClinGen.

**Table 1.** Associations between the susceptibility genes and breast cancer in 6 genetic resources

| Gene | Genetic Resources |  |  |  |  |  |
| --- | --- | --- | --- | --- | --- | --- |
|  | ClinGen | NCCN | OMIM | GHR | GeneCards | Gene-NCBI |
| *ATM* | Definitive | Increase | 1 | 1 | 1 | 1 |
| *BARD1* | Definitive | Potential increase | 1 | 1 | 1 | 1 |
| *BRCA1* | Definitive | Increase | 1 | 1 | 1 | 1 |
| *BRCA2* | Definitive | Increase | 1 | 1 | 1 | 1 |
| *CDH1* | Definitive | Increase | 1 | 1 | 1 | 1 |
| *CHEK2* | Definitive | Increase | 1 | 1 | 1 | 1 |
| *NBN* | Limited | Increase |  | 1 |  |  |
| *NF1* |  | Increase |  |  |  |  |
| *PALB2* | Definitive | Increase | 1 |  | 1 |  |
| *PTEN* | Definitive | Increase |  | 1 |  |  |
| *RECQL* | Moderate |  |  |  |  |  |
| *STK11* | Definitive | Increase | 1 | 1 |  |  |
| *TP53* | Definitive | Increase |  | 1 |  | 1 |

Abbreviations: GHR, GeneticsHomeReference

### 2. Diseases associated with BCSGs

There were 66 diseases initially identified, of which 40 diseases were confirmed to be associated with BCSGs by our evaluation (**Supplementary Table 3**). Besides breast cancer, malignant diseases including prostate cancer, pancreatic cancer, colorectal cancer, brain tumor, gastric cancer, ovarian cancer, and sarcoma were associated with at least 3 BCSGs (range: 3 to 8). However, *BARD1* and *RECQL* were only associated with breast cancer, without increased risk for any other diseases. Notably, *BRCA1/2, CHEK2*, and *PALB2* were also confirmed to be associated with male breast cancer.

**Table 2.**
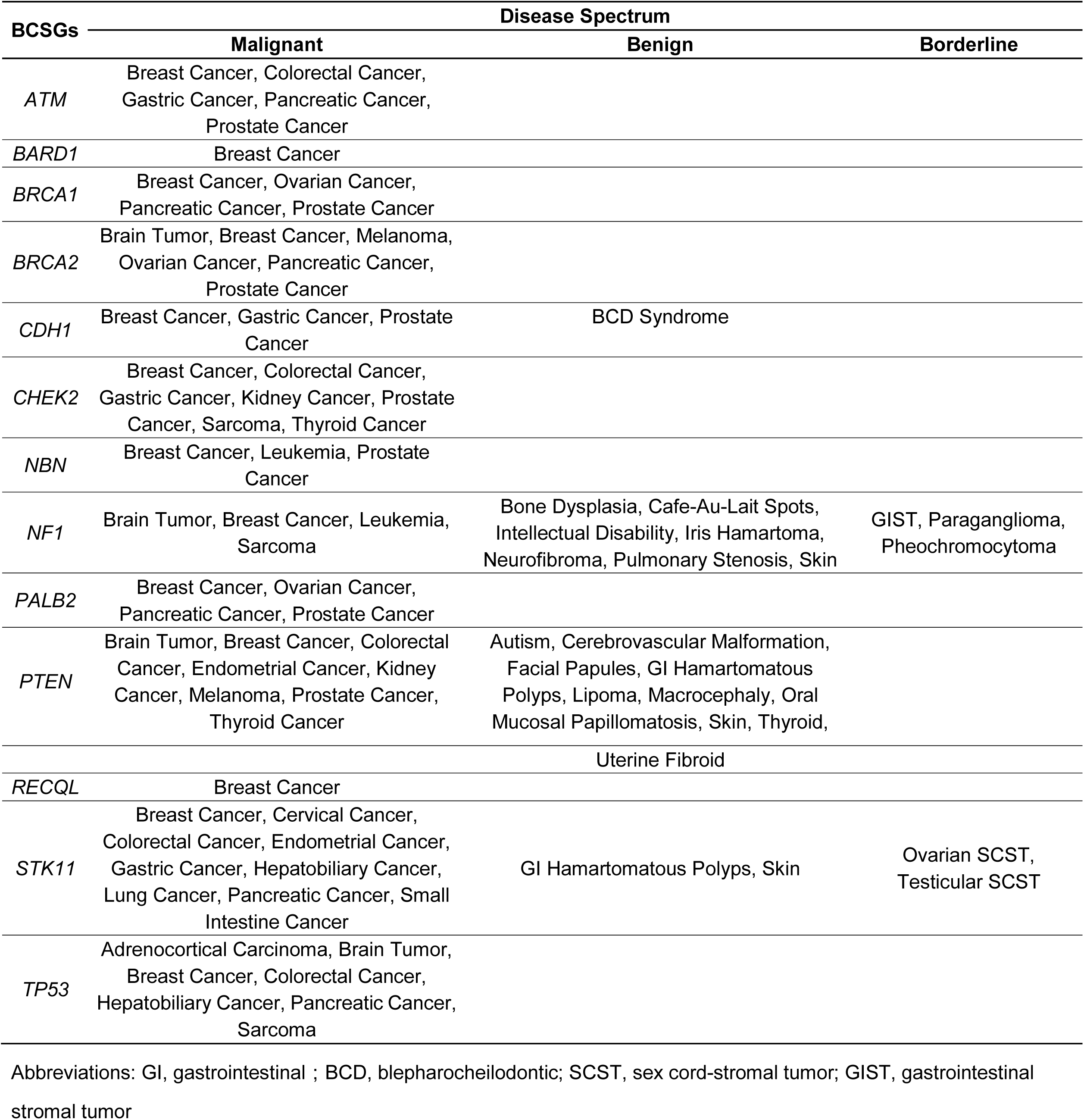
Diseases associated with breast cancer susceptibility genes

Disease spectrum of each BCSGs was shown in **Table 2**. Furthermore, several BCSGs are associated with specific syndromes, such as *NF1* with Neurofibromatosis Type 1, *PTEN* with Cowden Syndrome, *STK11* with Peutz-Jeghers Syndrome, and *TP53* with Li-Fraumeni Syndrome. The most common cancers of the syndromes were confirmed to be associated with the corresponding susceptibility genes by our procedure. For instance, Li-Fraumeni Syndrome-related cancers including sarcoma, brain tumor, breast cancer, adrenocortical carcinoma, and colorectal cancer were confirmed to be associated with *TP53*.

### 3. Disease spectrum of BCSGs and the corresponding resources

There were 171 gene-disease associations initially identified in the six genetic resources and literature (**Supplementary Table 1**). As shown in **Figure 2**, a total of 87 gene-disease associations were confirmed by our evaluation. Among them, 85% (74/87) of gene-disease associations were confirmed by ClinGen and/or NCCN. Conversely, 9 gene-disease associations were absent from both ClinGen and NCCN but confirmed in 3 or more of the other four genetic resources, these include BRCA2-Brain Tumor, *CDH1-*Blepharocheilodontic (BCD) Syndrome, and NF1-Leukemia which were confirmed by the other 4 resources. Notably, 4 gene-disease associations, namely, ATM-Gastric Cancer, *CHEK2*-Gastric Cancer, *CHEK2*-Kidney Cancer, and *CHEK2*-Thyroid Cancer, were confirmed only by the literature review using NLP.

## Discussion

Although hereditary breast cancer is mainly associated with *BRCA1/2* pathogenic variants, it may also be associated with germline mutations in other genes. Thus, multi-gene panels usually include both high- and moderate-penetrance genes associated with breast cancer [8,26,27]. The 13 BCSGs included in our study are those previously established by ClinGen and NCCN. To outline the disease spectrum for the 13 BCSGs, we examined 6 reliable genetic resources combined with a literature review using natural language processing (NLP). Finally, 40 unique diseases were confirmed to be associated with the 13 BCSGs.

One of the authoritative resources used for this study is the NIH-funded ClinGen, which, in contrast to “expert panel” consensus assessments used by NCCN, creates a framework that provides evidence for the strength of the association between a gene and a disease risk through semi-quantitative classification [28]. The ClinGen classification is based on genetic evidence including case-level data and case-control data, as well as experimental evidence. The other authoritative resource employed for this study are the NCCN Guidelines—the recognized standard for clinical practice in cancer care using its frequently updated set of clinical practice guidelines. More than 1,300 physicians and oncology researchers from the NCCN Member Institutions comprise the expert panels. Hence, the gene-disease association was assigned ‘confirmed’ in our study if it was established by either ClinGen or NCCN. Although the standardized literature review method used by ClinGen is outstanding [11], this approach is time-consuming and leads to delay in reflecting the most recent findings. In addition, the gene-cancer associations listed on the NCCN guidelines may not comprehensive. Therefore, it is necessary to include other genetic resources and find associations missed or not yet addressed by ClinGen and/or NCCN.

Four other genetic resources (OMIM, Genetics Home Reference, GeneCards and Gene-NCBI), are also considered reputable and contain a comprehensive compendium of relationships between phenotypes and genotypes. However, these resources lack the strict curation processes for evaluating strength of evidence utilized by ClinGen or the expert panels employed by NCCN. Therefore, we rated the level of evidence from these 4 resources as lower than ClinGen and NCCN, and the gene-disease association was assigned ‘confirmed’ only if it was established by at least 3 of these sources if the relationship was not found in ClinGen or NCCN. Meanwhile, we understand that the likely valid gene-disease associations we identified that were not present in ClinGen or NCCN, may be explained in part by the observation that those entities work in a slow and deliberate manner that might not yet have allowed a full review of all associations.

Forty unique diseases were confirmed to be associated with BCSGs by our procedure. Each BCSG was associated with at least 3 diseases except *BARD1* and *RECQL*, which were only associated with breast cancer. *BARD1* shares strong structural homology with BRCA1 and has been demonstrated to be involved in the cellular DNA repair process [29]. The association between breast cancer and mutations in the *BARD1* gene was first found in a large case-control study of 65,057 women with breast cancer [8], where the prevalence of *BARD1* mutations was 0.18%, significantly greater than the controls (OR 2.16, 95% CI 1.31-3.63, *P* < 0.05). On the other hand, *RECQL*, was first identified as a novel breast cancer susceptibility gene in 2015, by two independent research groups [30,31]. Bogdanova et al. compared 2596 breast cancer patients and 2132 healthy females from central Europe and indicated that *RECQL*\* c.1667_1667+3delAGTA could represent a moderate-risk breast cancer susceptibility allele [32]. A recent study found a moderate risk of breast cancer in African American women with *RECQL* mutation [33]. In addtion, *RECQL* is considered associated with hereditary breast carcinoma in ClinGen (gene-disease validity: moderate) (https://search.clinicalgenome.org/kb/genes/HGNC:9948). However, there is no high-quality penetrance study that showed statistical significance for additional diseases beyond breast cancer.

Generally speaking, the BCSGs are thought to affect female breast cancer risk, but some are also associated with male breast cancer (MBC). Tai et al. evaluated 97 men with breast cancer from 1939 families. The cumulative risk of breast cancer was higher in both *BRCA1* and *BRCA2* male heterozygotes compared to those without a *BRCA1/2* pathogenic variant at all ages. The relative risk of developing breast cancer peaks in the 30s and 40s [34]. Another study analyzed 321 families with *BRCA2* mutations both retrospectively and prospectively, suggesting a cumulative risk for male breast cancer up to age 80 of 8.9% [35]. Based on these data, NCCN guidelines recommend that men with a *BRCA1/2* pathogenic variant should receive a clinical breast exam at a young age [12].

Notably, we found that *CHEK2* and *PALB2* were also associated with male breast cancer in GeneCards. We confirmed these associations by literature review based on the NLP procedure, with the literature showing strong evidence in penetrance studies. The *CHEK2/1100delC*, a truncating variant, is present in 13.5% of individuals from families with male breast cancer (*P* = 0.00015), and results in an approximately tenfold increase of breast cancer risk in men [36]. A population-based study found the *CHEK2/1100delC* was present in 4.2% of unselected male breast cancer cases, more prevalent than the frequency of 1.1% in 1,692 controls (OR 4.1, 95% CI 1.2-14.3, *P* = 0.05) [37]. Recently, Yang et al. analyzed data from 524 families with *PALB2* pathogenic variants from 21 countries and found an association between *PALB2* and risk of male breast cancer (RR 7.34, 95% CI 1.28 to 42.18, *P* = 0.026) [38]. Additionally, Pritzlaff et al. reviewed 715 male breast cancer patients who underwent germline multi-gene panel testing and found that pathogenic variants in *CHEK2* (OR 3.7, *P* = 6.24 × 10^-24^) and *PALB2* (OR 6.6, *P* = 0.01) were both significantly associated with breast cancer risk in men [39].

In the present study, 85% of gene-disease associations were confirmed by ClinGen and/or NCCN, underscoring the credibility of these two major resources. Nevertheless, 9 gene-disease associations were not found in ClinGen or NCCN, but were instead identified in at least 3 of the other four genetic resources. Furthermore, these associations were similarly supported by published studies with strong evidence of the association, underscoring the reliability our review criteria (e.g., ≥3 resources).

Of note, 4 gene-disease associations, i.e., ATM-Gastric Cancer, CHEK2-Gastric Cancer, CHEK2-Kidney Cancer, and CHEK2-Thyroid Cancer, were not identified in any of the 6 resources, but were confirmed by the NLP aided literature review. In 2015, Helgason et al. reported a GWAS of gastric cancer in a European population, using information on 2,500 population-based gastric cancer cases and 205,652 controls. They found a new gastric cancer association with loss-of-function mutations in ATM (OR 4.74, *P* = 8.0 × 10^-12^) [40]. A recent study reported that *ATM* carriers were significantly associated with lower protein expression in five cancer types, including gastric cancer [41]. A *CHEK2* mutation was also identified to predispose to gastric cancer (OR 1.6, *P* = 0.004), in particular to young-onset cases (OR 2.1, *P* = 0.01) [21]. Additionally, Näslund-Koch et al. examined 86,975 individuals from the Copenhagen General Population Study. The age-and sex-adjusted hazard ratio for *CHEK2/1100delC* heterozygotes compared with noncarriers was 5.76 (95% CI 2.12-15.6) for gastric cancer, and 3.61 (95% CI 1.33-9.79) for kidney cancer [22]. Furthermore, a case-control study reported a *CHEK2* mutation in 15.6% of unselected patients with papillary thyroid cancer, compared to 6.0% in age- and sex-matched controls (OR 3.3, *P* < 0.0001) [42]. Another *CHEK2* variant, c.470C allele, was shown to increase the risk of papillary thyroid carcinoma in female patients by almost 13-fold (OR 12.81, *P* = 0.019) [43].

As the medical literature continues to grow exponentially, it takes more time and energy for clinicians to extract useful information precisely and quickly. NLP procedures to aid the literature review present a promising solution. These procedures are based on training a computational algorithm with many annotated examples to allow the computer to “learn” and “predict” the meaning of human language. Our previous study illustrates how to train and evaluate an NLP-based medical abstract classifier [24,25,44]. In 2016, we built our own clinical decision support tool for cancer susceptibility genes, called ASK2ME^TM^ [44,45]. This tool provides labs, researchers, and clinical experts with estimated cancer risk of germline pathogenic variants, including the disease spectrum for each susceptibility gene. This tool has been recommended as a resource in recent clinical practice guidelines [46].

The NCCN guidelines for considering risk-reducing mastectomy and breast MRI are well established for carriers of high-risk genes *(BRCA1, BRCA2, PALB2)*, and guidelines on annual mammogram with consideration of breast MRI are also established regarding carriers with moderate-risk genes *(ATM, CHEK2, NBN)* [12]. Although there are controversies regarding *NBN*, a high-quality meta-analysis showed a significantly increased risk of breast cancer in patients with the variant 657del5 (OR = 2.42, 95%CI 1.54-3.80) [47], which established *NBN* as a breast cancer susceptibility gene. Women with genes such as *TP53, CDH1, PTEN, STK11* and *NF1* may be managed according to established guidelines for the associated cancer predisposition syndrome. For instance, in Li-Fraumeni syndrome, annual whole-body MRI is advised in *TP53* pathogenic variant carriers [48]. More aggressive interventions may be recommended, such as consideration of prophylactic gastrectomy if a CDH1 mutation is found, even in the absence of gastric cancer in the family [49]. This necessitates that clinicians stay current with management guidelines and access reliable information resources to implement these updates effectively for their patients (e.g., resources such as ASK2ME could aid with this). Risks of other cancers for those BCSG carriers appear to be modestly elevated, but whether this should alter screening recommendations is unknown. For example, the risk of leukemia with *“TP53”* is 1.6 times as high as the general population, but as the general population risk of leukemia is 0.9%, that is an absolute risk of only 1.4% by age 85 [50]. Although a pathogenic mutation in *TP53* is statistically associated with leukemia, it would be hard to justify intensive screening or prevention measures. It is beyond the scope of this paper to identify the penetrance for each gene-disease association, but this will be the target of future work. Our proposed expansion of disease-gene association reporting will require clinicians to counsel patients appropriately about their risk of additional diseases and to refer them to genetic counselors or other specialists (e.g., neurologist, urologist).

Evaluation based on six genetic resources could result in omissions of some phenotypes associated with BCSGs. We attempted to lessen this effect by including a literature review as an additional step. Another limitation is that the strict criteria we set for gene-disease associations (e.g., confirmed by ClinGen/NCCN, or at least 3 genetic resources) could mean that some diseases are overlooked. By reviewing the literature using NLP, we reevaluated those uncertain gene-disease associations to lessen this effect as much as possible. Although the comprehensiveness of our data seems to be conducive to more individualized care, this raises the problem of absence of management guidelines for patients who carry such variants. Additionally, the clinical utility of identifying potential diseases in BCSG carriers may conflict with current cost-efficacy constraints (i.e., interpreting variants, genetic counseling, overdiagnoses, and resulting anxiety in patients). Of note, we are making assumptions based on the available evidence, and we recognize that these authoritative sources, such as ClinGen and NCCN guidelines, update periodically. Thus, this study represents a snapshot of current knowledge and understanding, rather than a definitive conclusion.

## Conclusions

To the best of our knowledge, this is the first study to collate the disease spectrum of BCSGs from multiple resources and make it available in a single resource. Notably, we developed an innovative assessment process based on six genetic resources and literature review using an NLP procedure. Throughout our evaluation process, we accept that frequent updates of the disease spectrum will be necessary to adjust for new data in these genetic resources. Our study provides a reference point for future studies, showing that BCSG mutation carriers should also be cautious of other diseases beyond breast cancer, and highlights the necessity of broadening the criteria of management and improving outcomes for at-risk individuals. These disease spectrums are available in our database ASK2ME^TM^ (https://ask2me.org/index.php), which is constantly updated. Ongoing research based on accurate estimates of cancer risk needs to be conducted in terms of appropriate management strategies.

### Ethics approval and consent to participate

We used public database with no patient data, and individual informed consent was waived.

### Consent for publication

Not applicable.

## Data Availability

N/A.

## Acknowledgements

The authors acknowledge Ann S. Adams for editorial and writing assistance.

---

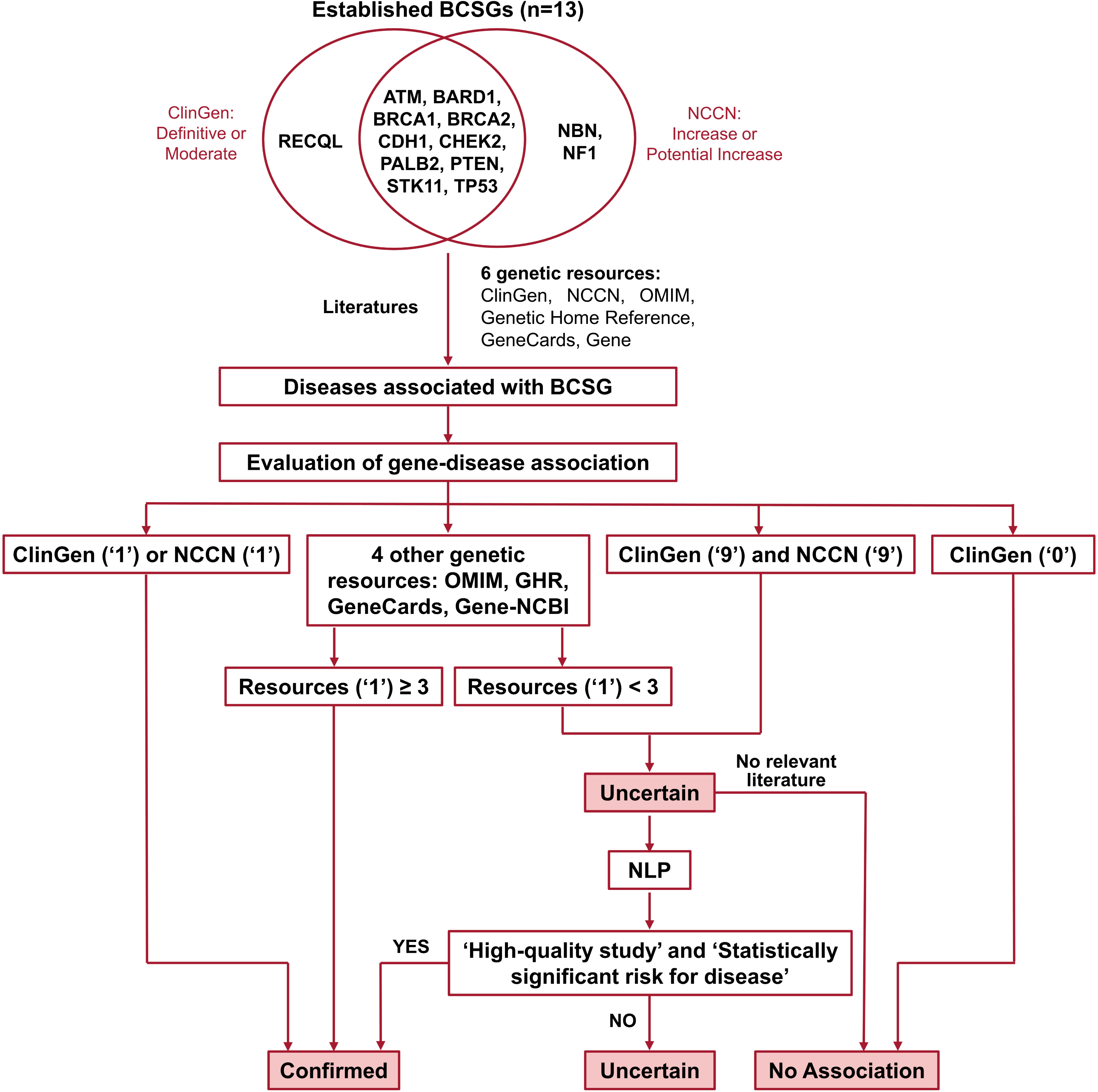

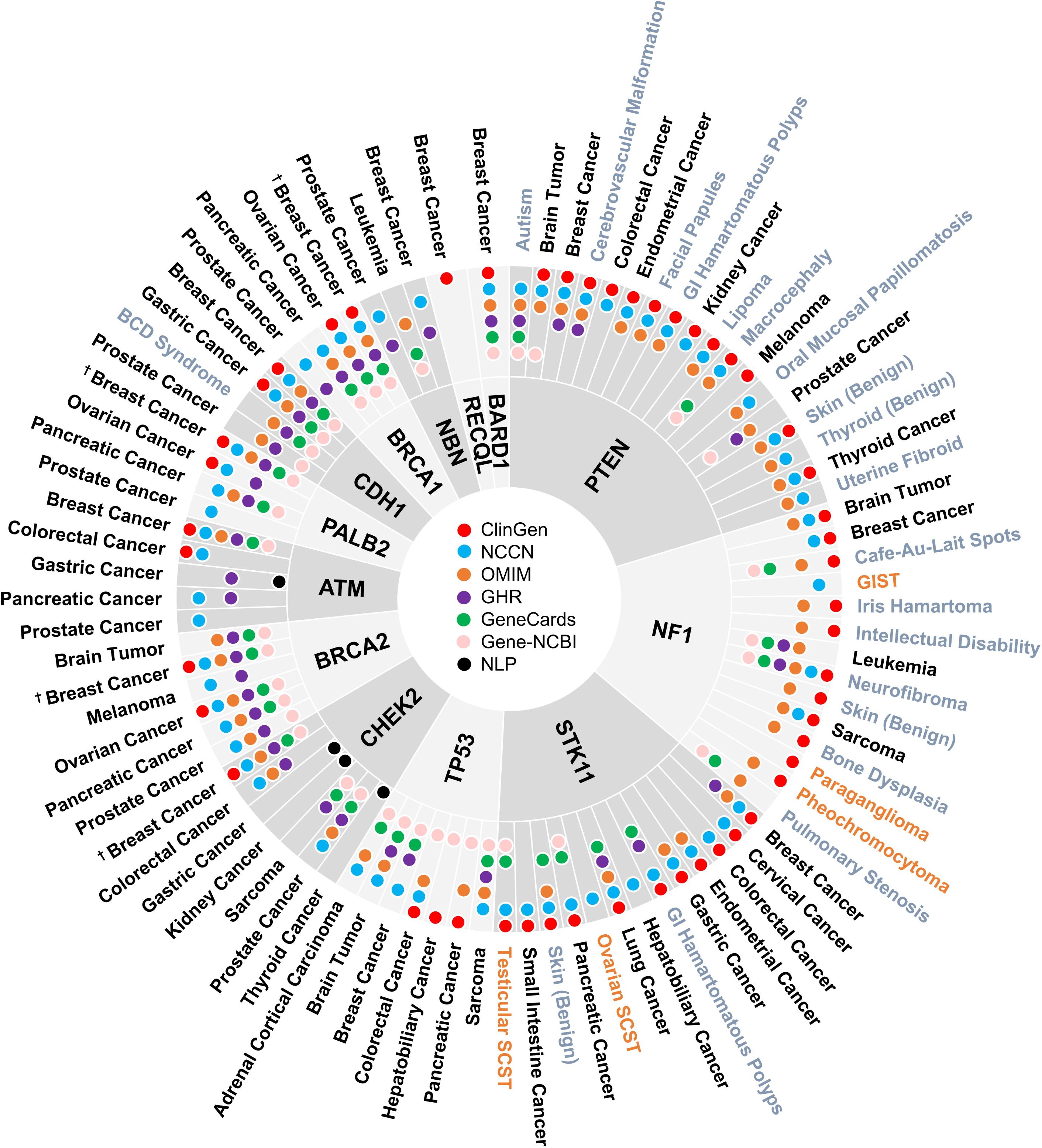

